# Factors influencing repeated decisions to decline cervical cancer screening among women living with HIV in Jos, Nigeria: a qualitative study

**DOI:** 10.64898/2026.04.22.26351475

**Authors:** Auwal Abubakar, Suraj Musa Inuwa, Maryam Jamila Ali, Abdullahi Mohammed Kabir, Yao Doe, Maiya G Block Ngaybe, Purnima Madhivanan, Jonah Musa

## Abstract

Women living with HIV face about a six-fold higher risk of cervical cancer, yet screening uptake remains low in many sub-Saharan African settings. We explored factors influencing repeated decisions to decline cervical cancer screening during routine HIV care among women living with HIV at a large HIV clinic in Jos, Nigeria.

Between September and December 2024, we conducted an exploratory qualitative study at the AIDS Prevention Initiative in Nigeria Clinic in Jos, Nigeria. We purposively recruited 27 women living with HIV aged 21 to 65 years who had never undergone cervical cancer screening and had repeatedly declined screening offers during routine HIV care, including at the current clinic visit. Semi-structured in-depth interviews were conducted in English or Hausa, audio-recorded, transcribed verbatim, and translated into English where needed. Data were analyzed thematically using theory-informed coding based on the Health Belief Model and Social Ecological Model.

Among 27 women living with HIV who had repeatedly declined screening, perceived susceptibility was often low or uncertain despite recognition of cervical cancer severity. Perceived benefits were acknowledged but were frequently outweighed by overlapping barriers, including knowledge gaps and misinformation, indirect and downstream costs, emotional barriers, logistical constraints, clinic-flow and service-delivery barriers, and anticipated stigma. Education, reminders, and supportive clinic processes acted as cues to action, and most participants expressed willingness to screen in future.

Among women living with HIV at this clinic who repeatedly declined screening when it was offered, perceived benefits were often outweighed by multilevel barriers. Screening programs may integrate fear-reduction and stigma-sensitive counseling with practical service delivery improvements, including shorter waiting times, reduced indirect costs, predictable and streamlined clinic flow, and consistent provider invitations and reminders, while addressing misinformation through community-embedded, culturally tailored messaging. These strategies may improve screening uptake and support more equitable cervical cancer prevention for women living with HIV in similar HIV-care settings.

## INTRODUCTION

Cervical cancer remains a major global public health concern and is the fourth most common cancer among women.[1] In 2022, an estimated 660,000 new cases and 350,000 deaths occurred worldwide, with approximately 94% of these deaths in low- and middle-income countries (LMICs).[1] Nigeria exemplifies this disparity, where cervical cancer is one of the leading causes of cancer-related mortality among women, with an estimated 7,100 deaths in 2022.[2] These regional differences reflect major inequities driven by limited access to human papillomavirus (HPV) vaccination, cervical cancer screening, and treatment services.[3,4] In sub-Saharan Africa, this burden is further exacerbated by the high prevalence of HIV and the substantially increased risk of cervical cancer among women living with HIV (WLHIV).[5] In 2020, the World Health Assembly adopted the World Health Organization’s (WHO) global strategy to accelerate cervical cancer elimination (90–70–90 targets by 2030), underscoring the urgency of closing screening gaps in high-burden settings.[6,7]

WLHIV have an approximately six-fold higher risk of cervical cancer compared with women without HIV.[5] This elevated risk largely reflects HIV-related immunosuppression, which increases the likelihood of acquiring and sustaining persistent infection with oncogenic HPV.[8–10] In some high HIV prevalence countries in Southern Africa, WLHIV account for over 40% of cervical cancer cases.[5]

Cervical cancer screening is effective for detecting precancerous lesions and for the secondary prevention of invasive cervical cancer.[11] WHO recommends HPV DNA testing as the preferred primary screening test and, for WLHIV, regular screening starting at age 25 years and repeated every 3 to 5 years.[12] Where HPV testing is unavailable, cytology or visual inspection with acetic acid (VIA) every 3 years is suggested.[12] WHO also supports the use of provider-collected or self-collected samples for HPV DNA testing, and the 2023 update reinforces validated self-sampling approaches as a way to expand coverage among under-screened populations.[12,13] During the study period, Nigerian HIV guidelines recommended cervical cancer screening for WLHIV every 3 years and stated that WLHIV should be screened regardless of age.[14]

Despite these recommendations, screening uptake across sub-Saharan Africa remains low. A 2021 systematic review estimated cervical cancer screening uptake at 12.87% (95% CI, 10.20 to 15.54) among women in sub-Saharan Africa.[15] A 2025 umbrella review across Africa reported pooled cervical cancer screening uptake of 19% (95% CI, 12 to 25).[16] Among WLHIV, a 2023 review estimated that about 30% (95% CI, 19 to 41) had ever been screened.[17] In Nigeria, uptake among WLHIV remains suboptimal. For example, in a cross-sectional study conducted in Jos, 81.6% had no prior evidence of Pap testing despite receiving HIV care.[18] In Plateau State, HIV prevalence among adults aged 15 to 64 years was 1.6% (95% CI, 1.2 to 2.0) in the 2018 Nigeria HIV/AIDS Indicator and Impact Survey (NAIIS).[19] The national adult prevalence was 1.5% (95% CI, 1.4 to 1.6).[20] Barriers persist, including limited awareness, stigma, and health system constraints.[21,22] Programmatic data from public health facilities in three Nigerian states reported HPV prevalence of about 16% among WLHIV.[23] Among women with a positive HPV result, only about 21% completed triage, highlighting substantial attrition in the multi-visit pathway.[23]

Although barriers to cervical cancer screening have been widely documented in sub-Saharan Africa and other LMIC settings,[17,21,24] fewer studies have examined how multilevel influences shape repeated decisions to decline screening among WLHIV in specific African settings. To our knowledge, published evidence from Plateau State remains limited on how stigma and service-delivery factors shape repeated decisions to decline cervical cancer screening among WLHIV. The Health Belief Model (HBM) is well suited to capture intrapersonal determinants (perceived susceptibility, perceived severity, perceived benefits, perceived barriers, cues to action, and self-efficacy),[25,26] while the Social Ecological Model (SEM) extends to interpersonal, institutional, and community contexts.[27] Accordingly, guided by HBM and SEM, we sought to understand factors influencing repeated decisions to decline cervical cancer screening offers through qualitative in-depth interviews with eligible WLHIV receiving HIV care at the AIDS Prevention Initiative in Nigeria (APIN) Clinic in Jos, Plateau State, Nigeria.

## METHODS

### Study design

We conducted an exploratory qualitative study using semi-structured, in-depth interviews to identify factors influencing repeated decisions to decline cervical cancer screening among eligible WLHIV. Repeated decline was defined as declining screening at the index visit and at least one prior routine HIV clinic visit, as documented in the screening register. Expressed intention-to-screen (defined as an explicit statement of willingness or plan to undergo screening now or at a future visit) was documented at the time of interview and summarized descriptively.

Analyses were guided by the HBM and the SEM. We used HBM constructs to organize intrapersonal themes and SEM levels to organize interpersonal, institutional, and community-level themes. We did not apply the SEM intrapersonal domain to avoid overlap with HBM constructs. SEM levels were operationalized from participants’ perspectives. This study is reported in accordance with the Consolidated criteria for Reporting Qualitative Research (COREQ) (S1 Checklist).[28]

### Study setting

The study took place at the APIN Clinic in Jos, an urban center in north-central Nigeria, between September and December 2024. APIN is a large HIV treatment facility in Plateau State and provides comprehensive HIV care, including routine cervical cancer screening for WLHIV. Its integrated screening services, large WLHIV cohort, and regular health talks provided a suitable context to examine reasons women declined screening.

During the study period, cervical cancer screening was integrated into routine HIV clinic flow and offered immediately after the standard health talks. Screening was conducted in an adjacent building to the HIV clinic and was provided at no cost to WLHIV. The screening methods routinely offered were visual inspection with acetic acid (VIA) and Pap smear (cervical cytology), delivered by nurses. VIA results were available immediately. Pap smear specimens were processed in the laboratory and interpreted by the Pathology unit, with results typically available within two weeks. WLHIV with abnormal screening findings were managed according to clinic protocol and referred for colposcopy when indicated.

### Participant recruitment and sampling

During routine HIV clinic sessions, nurses delivered standard health talks and offered cervical cancer screening to eligible WLHIV. WLHIV who declined the offer at the index visit and on ≥1 prior visit were identified using the screening register and were approached privately by trained research assistants, with assurances that participation was voluntary and unrelated to clinical care. In this study, participants comprised eligible WLHIV who had not previously undergone cervical cancer screening and who had repeatedly declined screening when it was offered during routine HIV care. Participant recruitment occurred from September 9 to December 12, 2024.

Those expressing interest underwent a brief study eligibility assessment and provided permission to be contacted. Interviews were scheduled at dates and times convenient for participants. We used purposive sampling to identify women who met the study criteria, and all eligible women identified during the study period consented and were interviewed (n = 27). For this study, eligibility was restricted to women aged 21 to 65 years. The lower age bound of 21 years was prespecified when the study was designed and was informed by HIV-specific cervical screening guidance available at that time.[29] WLHIV older than 65 years were not included in this study.

### Eligibility and exclusion criteria

#### Inclusion

Participants were eligible if they had a confirmed HIV diagnosis, resided in Plateau State, were aged 21 to 65 years, had not previously undergone cervical cancer screening, and had declined the offer of cervical cancer screening during routine HIV visits at the index visit and at least one prior visit. Participants also needed the capacity to provide informed consent.

#### Exclusion

Participants were excluded if they did not have an HIV diagnosis, accepted cervical cancer screening at the index visit, had cognitive impairment or serious illness that precluded participation, or were unable to communicate in English or Hausa.

### Data collection

Interviews were conducted in private consultation rooms at the APIN Clinic in Jos, Nigeria, between September and December 2024. Only the interviewer and the participant were present.

Each semi-structured interview was conducted in English or Hausa based on participant preference, lasted 30 to 60 minutes, and was audio recorded with consent.

The semi-structured interview guide was developed using HBM and SEM constructs and was reviewed for clarity, relevance, and cultural appropriateness. The guide was piloted with five WLHIV, and the pilot interviews were excluded from analysis. These five pilot participants were separate from the 30 women assessed for eligibility during the main study period and are not included in the analytic sample. Piloting led to minor refinements in the wording of probes and in the order of questions, while the core items remained unchanged. The final guide in English and Hausa covered sociodemographic characteristics, HBM domains including susceptibility, severity, benefits, barriers, cues to action, and self-efficacy, and SEM domains including interpersonal, institutional, and community influences.

No repeat interviews were conducted. No field notes were taken. Transcripts were not returned for participant review because written transcript review was not considered appropriate for all participants, given varied levels of formal education and the translation of Hausa-language interviews into English for analysis. Participants received reimbursement for time and transportation costs, ranging from ₦10,000 to ₦15,000 depending on travel distance and transport costs.

### Data management, translation, and analysis

Audio recordings were transcribed verbatim and de-identified. We assigned unique study IDs to each transcript. Hausa-language interviews were transcribed and then translated into English by a bilingual interviewer and the principal investigator. Both were native Hausa speakers and familiar with relevant medical terminology. Any discrepancies were resolved through discussion until agreement was reached. Data were stored on a password-protected external drive and in a secure University of Arizona Box folder, with access restricted to the study team. Quotations were presented verbatim, with light copy editing for readability. We used square brackets […] to indicate brief clarifications and ellipses (…) to indicate abridgment without altering meaning.

We conducted a combined deductive and inductive thematic analysis in ATLAS.ti (version 25.0.1), using Braun and Clarke’s six-phase framework to guide coding and theme development.[30] The initial codebook was primarily deductive and was informed by the HBM and the SEM. The codebook was expanded inductively based on participants’ narratives. Two members of the research team who were trained in qualitative methods independently coded all transcripts and resolved differences through discussion until agreement was reached. To enhance credibility and dependability, we used independent coding, discussion-based consensus, iterative refinement of the codebook, and verbatim quotations to anchor interpretation in participants’ accounts. Themes were iteratively reviewed and named, then mapped to HBM and SEM domains. We monitored thematic redundancy during iterative coding of successive interviews and judged by the end of data collection that no substantively new themes were emerging. All eligible women identified during the study period were interviewed (n = 27). For reporting, themes were organized by HBM constructs and by SEM levels and presented with participant-level counts (n/N). These counts are descriptive and are presented to indicate the breadth of subthemes across interviews. Theme counts were calculated at the participant level, and participants could contribute to more than one theme. Where useful, we report the number of participants who mentioned a subtheme to indicate breadth across interviews. Unless otherwise specified, denominators exclude missing or unknown values. In the narrative, we use qualitative frequency terms as shorthand. We define “few” (less than 25% of participants), “some” (25% to 49%), “many” (50% to 74%), and “most” (75% or more).

### Researcher reflexivity

Interviews were conducted by the principal investigator, AA, a male physician and public-health researcher with postgraduate training in public health (MBBS, MPH, MSc, DrPH) and formal training in qualitative methods, and three female research assistants, including one nurse employed at the APIN Clinic. All interviewers received study-specific training in qualitative interviewing. Apart from the clinic nurse interviewer, no prior relationship existed between the principal investigator or the other research assistants and participants. The nurse interviewer conducted interviews in a research capacity and may have been known to some participants through her work at the clinic. During the consent process, participants were informed that the interview team was conducting the study independently of routine clinical decision-making and that the purpose of the study was to understand women’s views and experiences of cervical cancer screening. The principal investigator has an interest in improving cervical cancer screening among WLHIV and was aware that this perspective could shape expectations about the value of screening and the interpretation of participants’ accounts.

### Ethical considerations

Ethical approval was obtained from the Jos University Teaching Hospital Health Research Ethics Committee (JUTH HREC; Ref: JUTH/DCS/IREC/127/XXXI/2711; National Health Research Ethics Committee (NHREC) registration number: NHREC/JUTH/05/10/22). The University of Arizona Institutional Review Board acknowledged the study under an external IRB reliance arrangement (IRB Submission ID STUDY00005623). Written informed consent was obtained from all participants, including consent for audio recording of interviews.

## RESULTS

We assessed 30 WLHIV for eligibility. Three women were ineligible because they were aged 18 to 20 years, below the study eligibility age. Twenty-seven participants completed interviews (n = 27). All eligible women who were approached consented to participate, and none withdrew. Although all 27 declined screening at the index visit, all participants had also declined screening on at least one prior routine HIV clinic visit, as documented in the screening register. At the time of interview, 25/27 (92.6%) participants expressed willingness or a plan to undergo cervical cancer screening now or at a future visit, despite declining screening at the index visit.

### Sociodemographic characteristics

Among the 26 participants with known age, the mean age was 45.5 years (standard deviation [SD], 9.6; range, 27 to 65). Many were married (19/27, 70.4%) and self-employed (16/27, 59.3%). Education ranged from no formal education (5/27, 18.5%) to tertiary education (9/27, 33.3%). Income reporting was limited, with 13/27 (48.1%) unable to estimate annual income. Among those who provided a household income estimate (n = 14, including those reporting no income), the median household income was ₦250,000 per year (interquartile range [IQR], ₦100,000–₦875,000). Complete category counts are in Table 1.

**Table 1.**
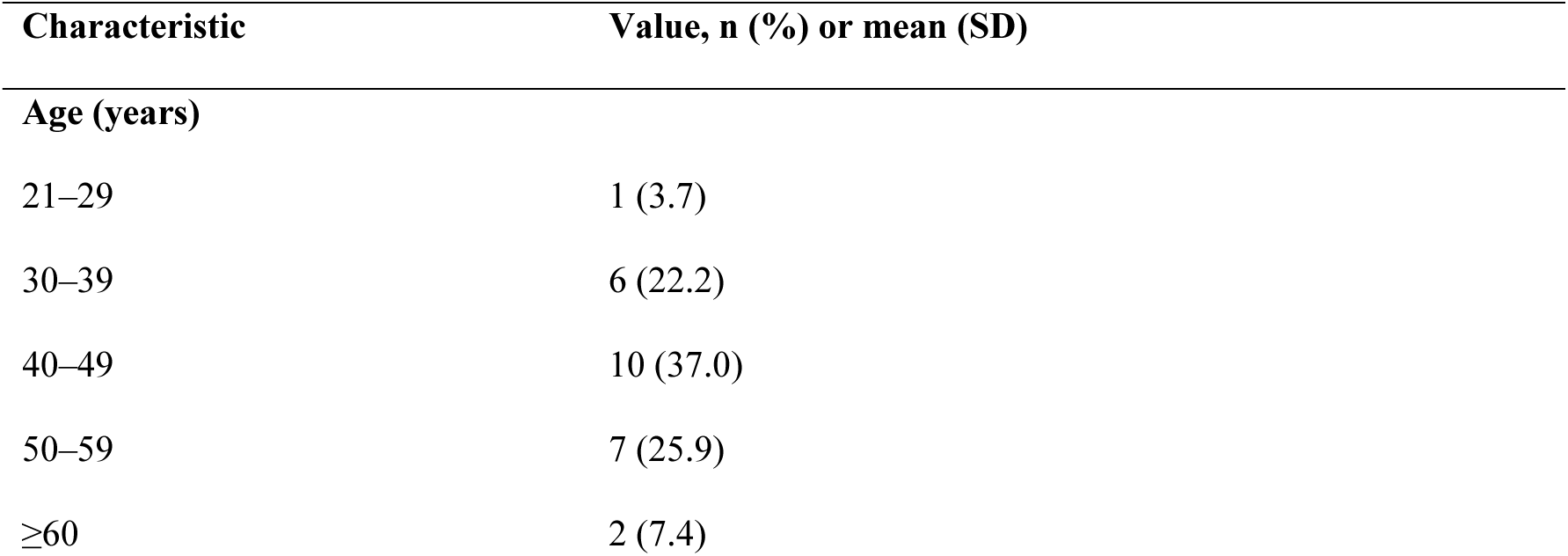
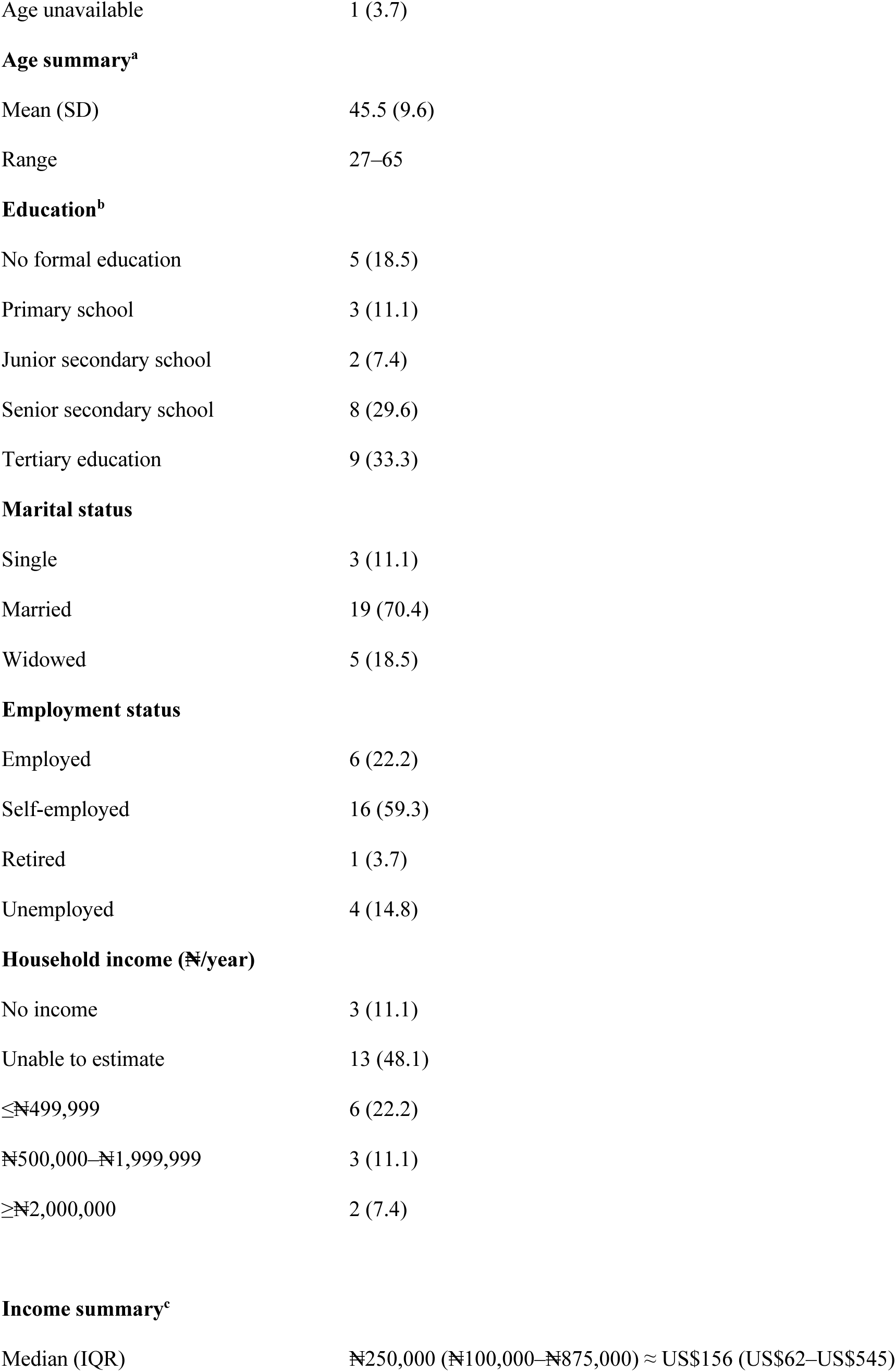
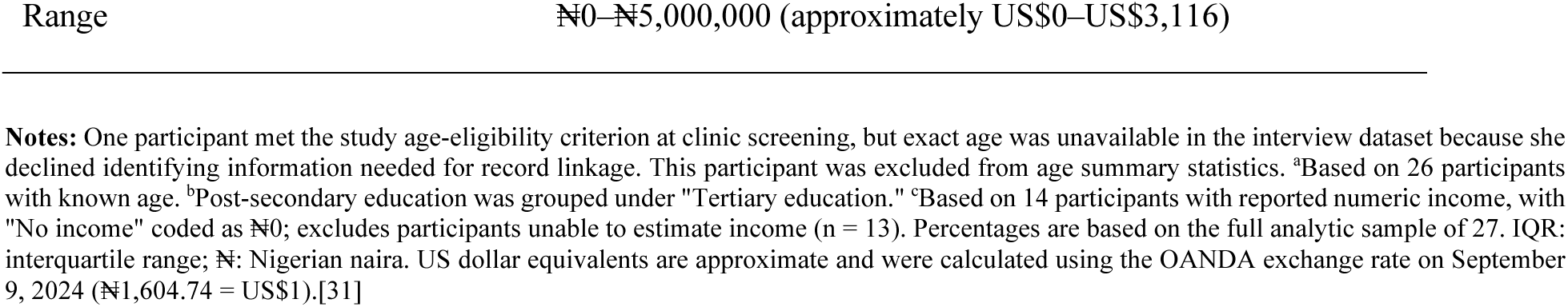
Sociodemographic characteristics of WLHIV who declined cervical cancer screening (N = 27)

### HBM

We organized codes under HBM constructs (perceived susceptibility, perceived severity, perceived benefits, perceived barriers, cues to action, and self-efficacy). Subthemes, participant-level counts, and illustrative quotations are presented in Table 2. The accompanying narrative provides a brief synthesis across constructs.

**Table 2.**
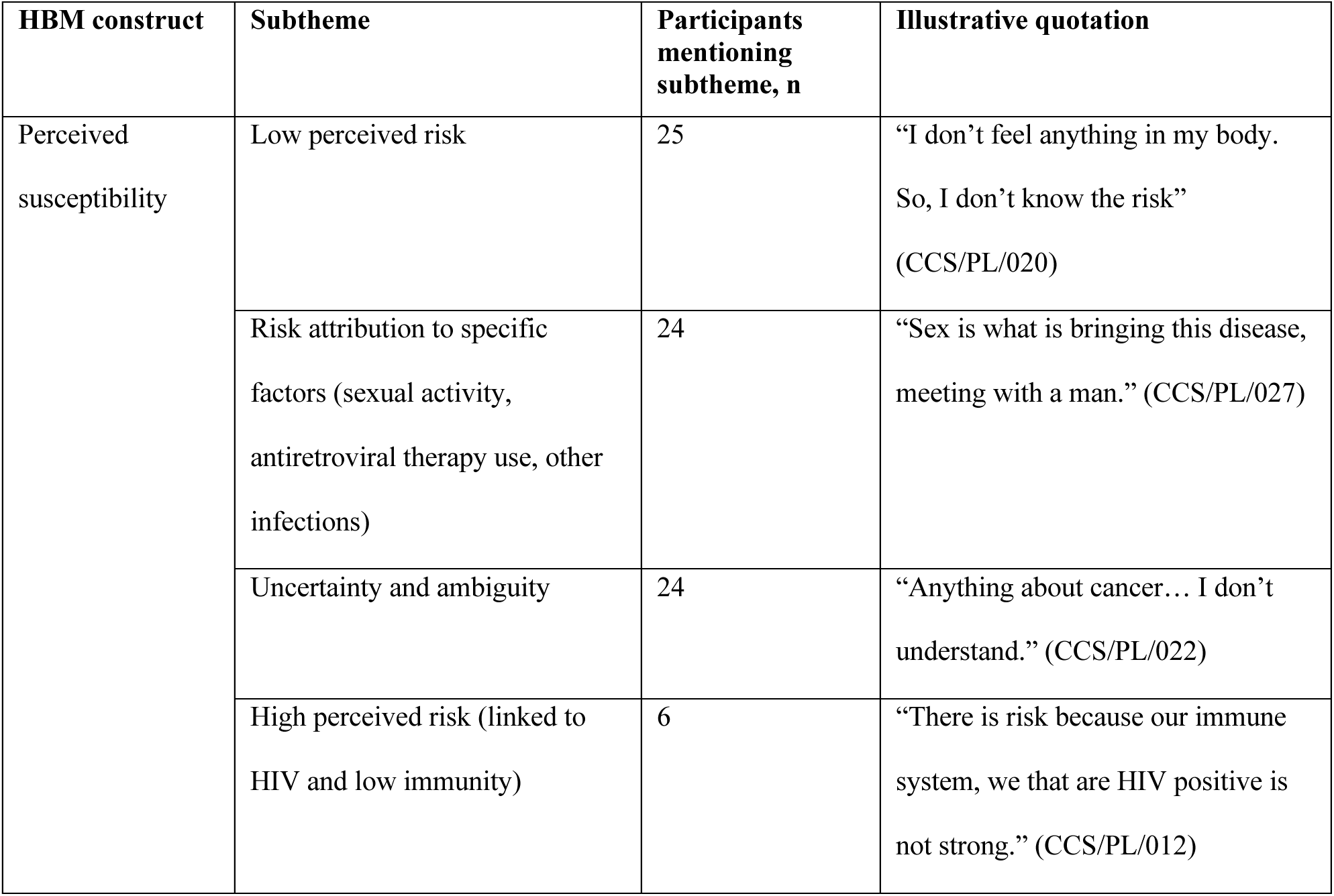
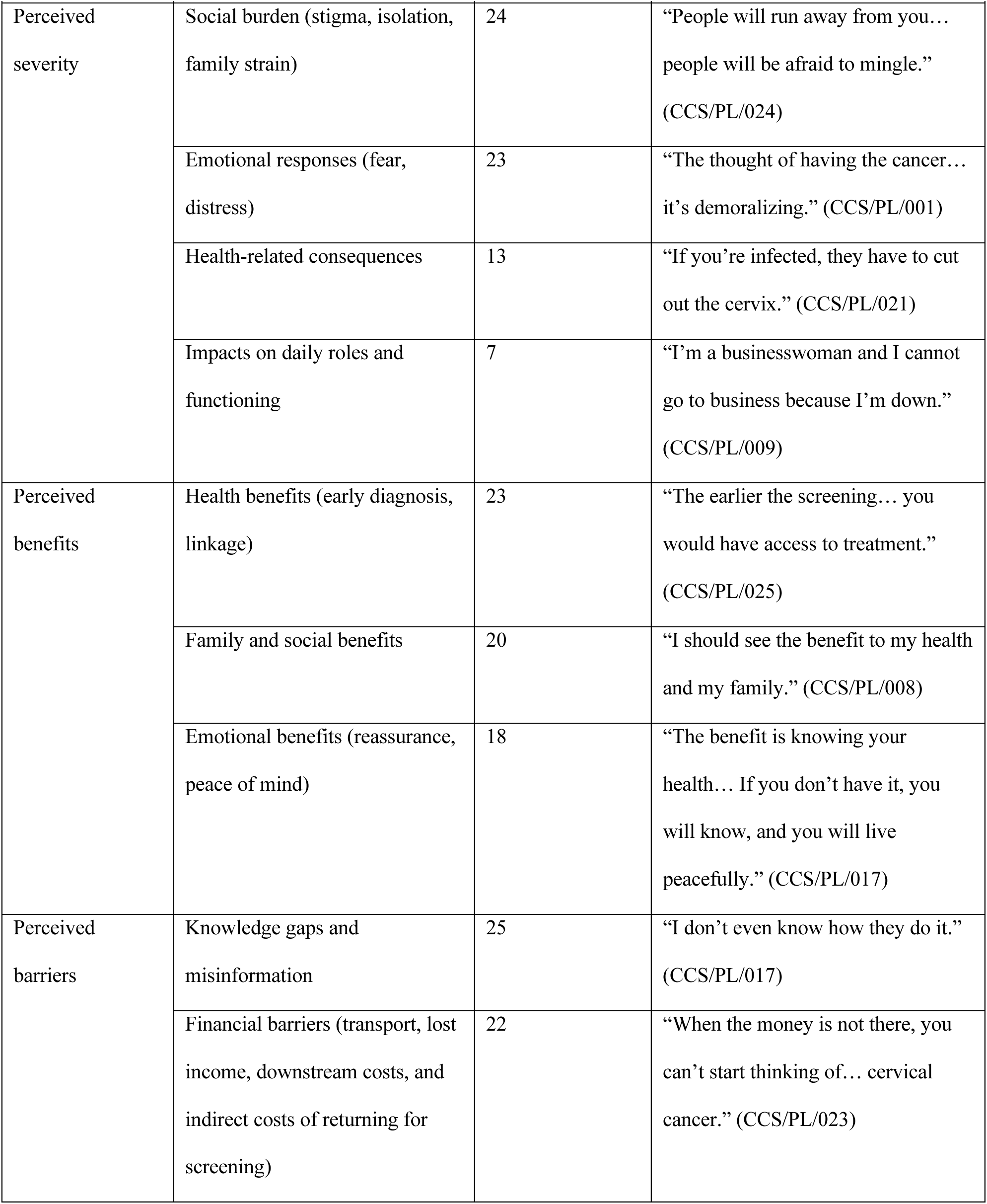
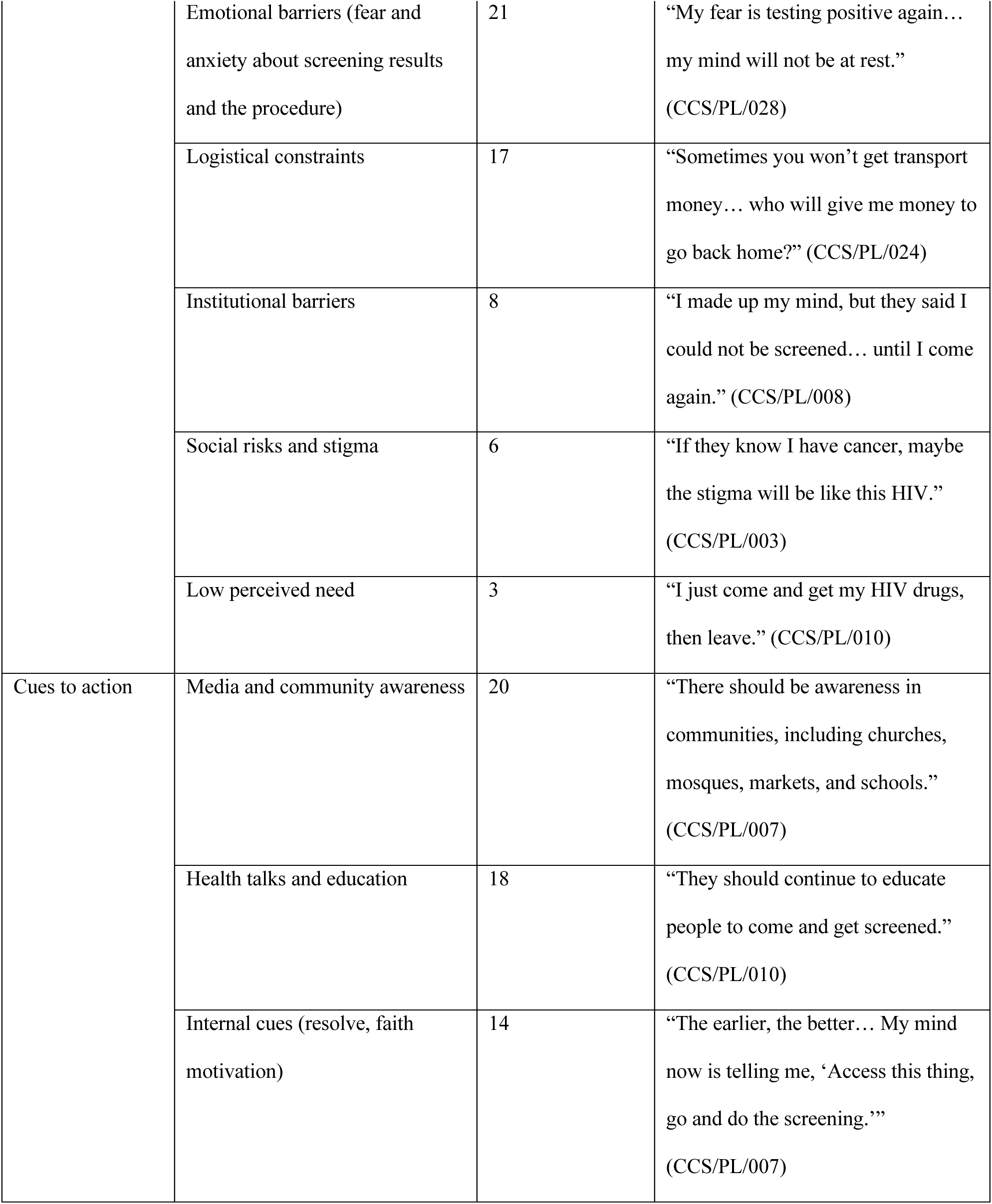
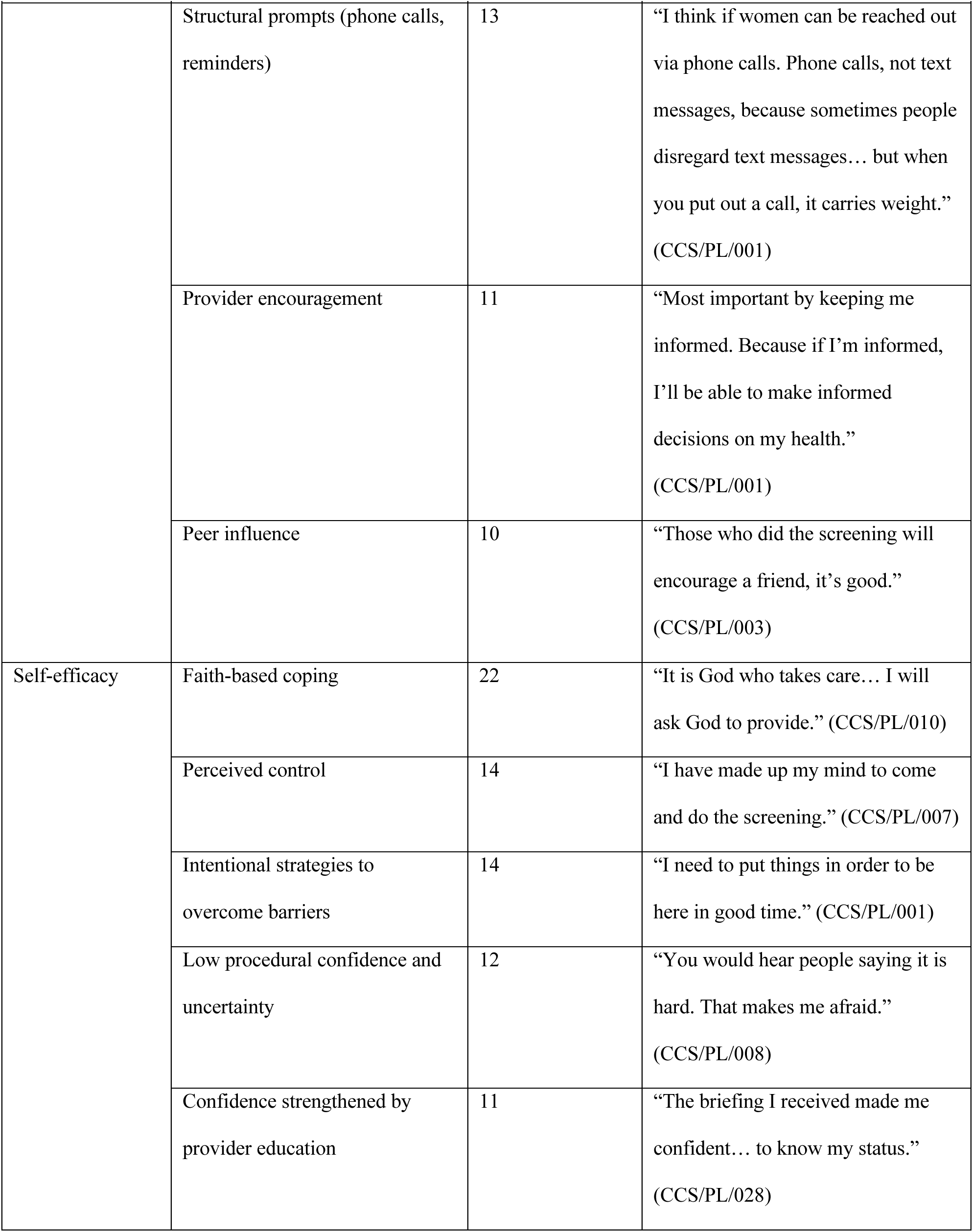
HBM-derived themes, subthemes, and illustrative quotations among WLHIV who declined screening (N = 27)

| HBM construct | Subtheme | Participants mentioning subtheme, n | Illustrative quotation |
| --- | --- | --- | --- |
| Perceived susceptibility | Low perceived risk | 25 | "I don't feel anything in my body. So, I don't know the risk" (CCS/PL/020) |
|  | Risk attribution to specific factors (sexual activity, antiretroviral therapy use, other infections) | 24 | "Sex is what is bringing this disease, meeting with a man." (CCS/PL/027) |
|  | Uncertainty and ambiguity | 24 | "Anything about cancer... I don't understand." (CCS/PL/022) |
|  | High perceived risk (linked to HIV and low immunity) | 6 | "There is risk because our immune system, we that are HIV positive is not strong." (CCS/PL/012) |

| **HBM construct** | **Subtheme** | **Participants mentioning subtheme, n** | **Illustrative quotation** |
| --- | --- | --- | --- |
| Perceived severity | Social burden (stigma, isolation, family strain) | 24 | “People will run away from you... people will be afraid to mingle.” (CCS/PL/024) |
|  | Emotional responses (fear, distress) | 23 | “The thought of having the cancer... it’s demoralizing.” (CCS/PL/001) |
|  | Health-related consequences | 13 | “If you’re infected, they have to cut out the cervix.” (CCS/PL/021) |
|  | Impacts on daily roles and functioning | 7 | “I’m a businesswoman and I cannot go to business because I’m down.” (CCS/PL/009) |
| Perceived benefits | Health benefits (early diagnosis, linkage) | 23 | “The earlier the screening... you would have access to treatment.” (CCS/PL/025) |
|  | Family and social benefits | 20 | “I should see the benefit to my health and my family.” (CCS/PL/008) |
|  | Emotional benefits (reassurance, peace of mind) | 18 | “The benefit is knowing your health... If you don’t have it, you will know, and you will live peacefully.” (CCS/PL/017) |
| Perceived barriers | Knowledge gaps and misinformation | 25 | “I don’t even know how they do it.” (CCS/PL/017) |
|  | Financial barriers (transport, lost income, downstream costs, and indirect costs of returning for screening) | 22 | “When the money is not there, you can’t start thinking of... cervical cancer.” (CCS/PL/023) |
|  | Emotional barriers (fear and anxiety about screening results and the procedure) | 21 | “My fear is testing positive again... my mind will not be at rest.”<br>(CCS/PL/028) |
|  | Logistical constraints | 17 | “Sometimes you won’t get transport money... who will give me money to go back home?” (CCS/PL/024) |
|  | Institutional barriers | 8 | “I made up my mind, but they said I could not be screened... until I come again.” (CCS/PL/008) |
|  | Social risks and stigma | 6 | “If they know I have cancer, maybe the stigma will be like this HIV.”<br>(CCS/PL/003) |
|  | Low perceived need | 3 | “I just come and get my HIV drugs, then leave.” (CCS/PL/010) |
| Cues to action | Media and community awareness | 20 | “There should be awareness in communities, including churches, mosques, markets, and schools.”<br>(CCS/PL/007) |
|  | Health talks and education | 18 | “They should continue to educate people to come and get screened.”<br>(CCS/PL/010) |
|  | Internal cues (resolve, faith motivation) | 14 | “The earlier, the better... My mind now is telling me, ‘Access this thing, go and do the screening.’”<br>(CCS/PL/007) |
|  | Structural prompts (phone calls, reminders) | 13 | “I think if women can be reached out via phone calls. Phone calls, not text messages, because sometimes people disregard text messages... but when you put out a call, it carries weight.” (CCS/PL/001) |
|  | Provider encouragement | 11 | “Most important by keeping me informed. Because if I’m informed, I’ll be able to make informed decisions on my health.” (CCS/PL/001) |
|  | Peer influence | 10 | “Those who did the screening will encourage a friend, it’s good.” (CCS/PL/003) |
| Self-efficacy | Faith-based coping | 22 | “It is God who takes care... I will ask God to provide.” (CCS/PL/010) |
|  | Perceived control | 14 | “I have made up my mind to come and do the screening.” (CCS/PL/007) |
|  | Intentional strategies to overcome barriers | 14 | “I need to put things in order to be here in good time.” (CCS/PL/001) |
|  | Low procedural confidence and uncertainty | 12 | “You would hear people saying it is hard. That makes me afraid.” (CCS/PL/008) |
|  | Confidence strengthened by provider education | 11 | “The briefing I received made me confident... to know my status.” (CCS/PL/028) |

**Note:** n indicates the number of participants (out of N = 27) who mentioned the subtheme at least once; CCS stands for Cervical Cancer Screening and PL for Plateau. Participant identifiers are formatted as CCS/PL/###, where ### is a unique participant ID. Quotations are de-identified and lightly edited for brevity and clarity; ellipses indicate minor omissions; bracketed text clarifies meaning. Participant identifiers are non-sequential because study IDs were assigned at recruitment and retained after exclusion of ineligible cases.

HBM-derived subthemes and representative quotations are presented in Table 2. Risk appraisals were frequently symptom-based. Most women interpreted the absence of symptoms as evidence of low susceptibility *(“I don’t feel anything in my body. So, I don’t know the risk” (CCS/PL/020))*. Some viewed older age or periods of sexual inactivity as lowering the need for screening *(“the reason I did not bother myself is because I am old” (CCS/PL/006))*. Uncertainty about cervical cancer and misconceptions about risk factors or eligibility contributed to ambiguity and deferral, even among the minority who recognized heightened vulnerability related to HIV.

Participants commonly described cervical cancer as highly serious, but perceived severity did not consistently translate into same-day screening. In several accounts, high severity coexisted with fear, anticipated stigma, and concern about the affordability of care, and participants described these concerns alongside screening deferral *(“Truth be told, my main reason is fear… and the lack of finances to come” (CCS/PL/028))*. Concerns about downstream costs were explicit in some narratives *(“Treating it can be so expensive… and only the rich can afford it” (CCS/PL/028))*.

Participants articulated clear benefits of screening, including early detection, reassurance, and preserving family and social roles *(“if it is early detected, it can stop the spread” (CCS/PL/011))*. However, these perceived benefits were often outweighed at the point of offer by overlapping barriers. Women described barriers as interlinked, with limited procedural knowledge amplifying fear of results and anticipated discomfort *(“I don’t even know how they do it” (CCS/PL/017))*. In a few accounts, participants appeared to conflate screening with other preventive interventions, including vaccination, reflecting uncertainty about the purpose and process of cervical cancer screening *(“not knowing that they are helping us with the vaccine… The important thing is to know your health status” (CCS/PL/027))*. Other interconnected barriers included indirect costs, anticipated downstream costs, time constraints, caregiving responsibilities, and occasional service disruptions, resulting in index-visit non-initiation *(“I will rush, and I will not wait for the test” (CCS/PL/024); “When I came, they said that I could not be screened… until I came again” (CCS/PL/008))*. Some participants also described timing-related deferral during menstruation or pregnancy, and others described forgetting to proceed for screening after collecting HIV medications or leaving without being screened when the clinic visit became prolonged *(“anytime I came… I will be on my period… I usually come after six months to get my drugs… and I forget” (CCS/PL/004)).* Another participant described pregnancy, transfer to the children section, fatigue, and leaving without screening *(“I became pregnant then I withdrew… I was transferred there (children section) … you might be tired and time will go so I will just leave” (CCS/PL/012))*.

Women identified cues that could increase readiness to screen, including consistent clinic education, community-based awareness, reminder systems aligned with HIV appointments, and reduced indirect costs associated with attending screening *(“We should call them… just call them… they called me the morning” (CCS/PL/009))*. Self-efficacy was dynamic and appeared strengthened by provider explanations, faith-based coping, and practical planning to attend early *(“I need to put certain things in order… for me to always be here in good time” (CCS/PL/001); “after they gave a clear explanation… I realized there was nothing to fear, it’s not painful” (CCS/PL/021))*, but was contingent on predictable availability and a clear, streamlined screening process during routine HIV clinic visits *(“If we come to collect our drugs and decide we want to get screened… they said… I could not be screened… until I came again” (CCS/PL/008); “they should show you what to do… tell us where to follow… we should not be left with the result” (CCS/PL/012))*.

### SEM

Moving from individual beliefs to context, we summarize interpersonal, institutional, and community-level influences on barriers to screening uptake at the index visit. Subthemes, participant-level counts, and representative quotations are presented in Table 3. The narrative below synthesizes cross-cutting patterns across SEM levels.

**Table 3.** SEM-derived themes, participant-level counts, and illustrative quotations among WLHIV who declined screening (N = 27)

| SEM level | Subtheme | Participants mentioning subtheme, n | Illustrative quotation |
| --- | --- | --- | --- |
| Interpersonal | Interpersonal encouragement (relatives, partners or peers) | 24 | “My senior sister... advised me to come and do the screening.” (CCS/PL/004) |
|  | Shared fears within networks | 10 | “They think that if you have cancer, it means death... They’re also afraid of the financial problems that might come with it.” (CCS/PL/018) |
|  | Discouragement from close contacts | 9 | “If nothing is wrong with you, why go?” (CCS/PL/028) |
|  | Absence of dialogue about screening | 7 | “I have never sat with anyone to discuss... the screening.” (CCS/PL/006) |
|  | Limited influence and autonomy emphasized | 7 | “Their opinions didn’t affect my decision... I came here to be checked.” (CCS/PL/019) |
|  | Outward advocacy despite personal deferral | 6 | “I ask women if they’ve screened. If not, I tell them to come.” (CCS/PL/028) |
| Institutional | Institutional support mechanisms (reminders, health talks) | 20 | “Every clinic visit they remind us. We are the ones dodging.” (CCS/PL/003) |
|  | Quality of provider interaction | 20 | “They give us health talks... and they are ready to assist us.” (CCS/PL/025) |
|  | Affordability | 16 | “We’re grateful that it has been made free for us.” (CCS/PL/028) |
|  | Distance and transport constraints | 13 | “The screening services... are far from my home.” (CCS/PL/011) |
|  | Proximity and streamlined delivery | 9 | “The testing area is... very close to the clinic.” (CCS/PL/025) |
|  | Health system performance (waits, delays) | 5 | “When we are in the APIN Clinic, we will not finish on time... so I will rush, and I will not wait for the test.” (CCS/PL/024) |
|  | Navigation and information clarity | 5 | “They explain, but I don’t know where to go and get screened.” (CCS/PL/013) |
|  | Discouraging or unsupportive provider communication | 3 | “They should please talk to us calmly... Not with shouting like, ‘If you don’t do it, you’ll die,’ or ‘Your cervix will be removed!’ No, that just creates fear.” (CCS/PL/012) |
| Community | Religious and cultural context (often perceived as non-limiting) | 16 | "I told you nobody has a problem with it they only encourage doing it because it is good." (CCS/PL/004) |
|  | Stigma related to cervical cancer | 10 | "Cancer is often viewed... as a death sentence." (CCS/PL/011) |
|  | Community misconceptions | 9 | "People say screening is a waste of money." (CCS/PL/025) |
|  | Lack of community awareness | 8 | "People have never even heard about cervical cancer." (CCS/PL/024) |
|  | Community-based awareness programs | 7 | "Go into those places and enlighten the women." (CCS/PL/004) |
|  | Traditional remedies or spiritual explanations | 3 | "They believed you should use a leaf from the bush and rub it over the vagina." (CCS/PL/012) |

**Note:** n indicates the number of participants (out of N = 27) who mentioned the subtheme at least once; CCS stands for Cervical Cancer Screening and PL for Plateau. Participant identifiers are formatted as CCS/PL/###, where ### is a unique participant ID. Quotations are de-identified and lightly edited for brevity and clarity; ellipses indicate minor omissions; bracketed text clarifies meaning. Participant identifiers are non-sequential because study IDs were assigned at recruitment and retained after exclusion of ineligible cases.

#### Interpersonal Factors

At the interpersonal level, women described social relationships and support networks as both supportive and constraining. Encouragement from relatives, partners, peers, and support-group members could normalize screening and reduce fear *(“My senior sister… advised me to come and do the screening.” (CCS/PL/004))*. However, similar networks also conveyed anxiety, fatalistic interpretations of cancer, and discouraging messages that undermined intentions *(“If nothing is wrong with you, why go?” (CCS/PL/028))*. Some women reported limited discussion about screening or emphasized autonomy in decision-making.

Notably, a few described advising other women to screen despite personally deferring, showing that some women encouraged screening for others while still deferring screening themselves *(“I ask women if they’ve screened. If not, I tell them to come.” (CCS/PL/028))*.

#### Institutional Factors

Institutional influences centered on clinic delivery and access. Women described routine counseling, reminders, and respectful communication as facilitating readiness to screen during routine clinic visits *(“Every time we come to the clinic, they remind us to get screened. Honestly, we are the ones who have been dodging.” (CCS/PL/003))*. However, a minority perceived fear-based or threatening messaging as increasing anxiety and discouraging screening, and in these accounts, women emphasized the importance of calm, supportive counseling. As one woman advised, providers should avoid shouting messages. Clinic flow sometimes limited exposure to these educational cues. One participant noted that after being transferred to the children section, health talks were less consistent there *(“They don’t usually do the health talk as they do here” (CCS/PL/012))*. The ability to translate intention into same-day initiation was shaped by affordability and indirect costs, transport and distance, and the predictability and efficiency of service delivery. Co-location of screening with HIV care was perceived as reducing effort when navigation was clear. Conversely, long waiting times, service interruptions, delayed results, and uncertainty about where to go or what the procedure entails could deter completion during routine clinic visits *(“I was told the workers were not around” (CCS/PL/005); “They explain, but I don’t know where to go and get screened” (CCS/PL/013))*.

In addition, some women described leaving after collecting HIV medications without proceeding to screening *(“I just come and get my HIV drugs, then leave.” (CCS/PL/010))*.

#### Community-level Factors

At the community level, many women did not perceive religious or cultural norms as directly prohibitive *(“There is nothing that can affect my decision to get screened. In my community, there are no beliefs or norms affecting this…” (CCS/PL/029))*.

However, community narratives and misconceptions contributed to stigma, fear, and low awareness of screening *(“Cancer is often viewed… as a death sentence.” (CCS/PL/011))*.

Some participants referenced traditional remedies or spiritual explanations as alternative pathways that could delay biomedical care. Participants suggested that community-embedded education through faith venues, markets, schools, and primary health care settings could increase accurate knowledge and normalize screening *(“There should be awareness in communities, including churches, mosques, markets, and schools.” (CCS/PL/007); “Go into those places and enlighten the women.” (CCS/PL/004))*.

## DISCUSSION

We sought to understand behavioral and contextual factors underlying repeated decisions to decline cervical cancer screening among eligible WLHIV at the APIN Clinic in Jos, Plateau State. Guided by the HBM and the SEM, we found that perceived susceptibility was often low or ambiguous despite high perceived severity. Although benefits were recognized, emotional (fear of test results and anxiety about the screening procedure), financial, and logistical barriers frequently outweighed them, contributing to repeated deferral across clinic encounters, including the index visit.

HBM constructs operated in dynamic interplay with multilevel contexts mapped by SEM. Fear and anticipated stigma at interpersonal and community levels sometimes translated perceived severity into avoidance. In contrast, provider encouragement, reminders, respectful care and predictable availability of free screening services appeared to support readiness and stated future intention to screen. When these clinic-level supports were absent or inconsistent at the index visit, women continued to defer despite recognizing benefit. Fatalism and misinformation tended to amplify perceived severity while obscuring susceptibility, and stigma and cost narratives functioned as persistent perceived barriers. Participants described clinic talks, phone reminders, and peer testimonies as cues to action that could bolster self-efficacy.

Patterns in our data mirror evidence from Nigeria and other sub-Saharan African (SSA) settings, where fear of test results and anticipated stigma are well-documented barriers to cervical cancer screening uptake.[22,32–35] Other qualitative work further highlights modesty-related concerns, spousal approval, embarrassment about pelvic examinations, and preference for female providers as barriers reported in some settings, although these were not prominent in our sample.[22,32–35] Systematic reviews also underscore the role of financial and health system access barriers, including the cost of screening, transport costs, distance to facilities, and limited local availability of services, in reducing uptake across SSA.[15,21,32–34] Taken together, this literature indicates that these behavioral and structural barriers are recurrently reported across the region, rather than being unique to our study population.[15,21,32–35] The symptom-driven mental model, in which women linked risk to the presence of symptoms, reflects reports that women seek screening mainly when unwell, highlighting gaps in awareness of asymptomatic disease progression. Similar patterns, where low perceived risk and fear of positive results contribute to low screening uptake, have been reported among women in Nigeria and other sub-Saharan African settings.[32,34] Perceived benefits, including early detection and peace of mind, were evident, consistent with meta-analytic findings that greater knowledge about cervical cancer and screening is associated with higher screening uptake in sub-Saharan Africa.[15] However, these benefits were often overshadowed by emotional fears, financial constraints, and stigma. Although cervical cancer screening at APIN Clinic was offered at no cost to WLHIV, participants described financial barriers related to transport, time away from work, childcare, and concerns about potential costs of follow-up care. This tension mirrors prior reports that screening costs, fear of positive results, and stigma-related concerns can deter cervical cancer screening.[22,32,35] In our context, HIV-related stigma, community misconceptions, and low awareness appeared to compound cervical cancer fears and reinforce avoidance, consistent with SEM’s community-level influences and echoing calls for expanded community-based education on cervical cancer and screening.[33,36]

Clinic health talks, provider encouragement, and reminder systems appeared to strengthen readiness to screen,[37,38] but inconsistent community outreach and limited peer discussion remained missed opportunities. Self-efficacy varied as some women showed resilience (*“I’ve made up my mind to come and do the screening,” (CCS/PL/007*)), often linked to faith (*“Once I start praying, all the fear disappears,” (CCS/PL/004*)), but institutional barriers (*“I was told the workers were not around,” (CCS/PL/005)*) and negative provider interactions (*“Many times, the health workers’ attitude is discouraging” (CCS/PL/022)*) undermined this for others.

Together, these findings illustrate how a complex interplay of cognitive, emotional, social, and structural factors, often reinforcing one another, undermines screening participation among WLHIV despite available services. Understanding these layered dynamics is essential for designing responsive, stigma-sensitive interventions.

These findings have programmatic and policy implications for Nigeria and other high-burden settings, where the intersecting burdens of cervical cancer and HIV pose a complex public health challenge.[5,38,39] First, tailored education should address fear and stigma, reframing screening as empowering and clarifying asymptomatic disease. Second, community-based campaigns delivered through community venues and local-language outreach, with reminder systems tailored to participant preferences such as phone calls, can counter misconceptions (*“It would be good to go into those places and enlighten the women,” (CCS/PL/004)*) and may improve acceptability in low-resource Nigerian settings.[33] Third, clinic improvements could include shorter wait times, reliable staff availability, respectful and non-threatening communication, one-on-one explanation where needed, confidentiality-sensitive follow-up, and sustained no-cost screening. Together, these changes could strengthen self-efficacy and improve future uptake. These strategies are consistent with WHO guidance, which emphasizes integrated, accessible cervical cancer screening and treatment for WLHIV in high-burden settings.[12,38] Beyond program adjustments, our findings identify targets for mixed-methods verification, including provider interviews, client-flow time–motion studies, and policy and process review, to test and refine the most actionable levers.

This study has several limitations. First, because participants were identified at the point of care, findings reflect index-visit decisions, and some women may subsequently choose to screen.

Second, the single-site focus (APIN Clinic, Jos) may limit transferability to rural or non-clinic-attending WLHIV. The prespecified study age range of 21 to 65 years may limit transferability to women outside this age range, including younger and older women attending the clinic. Because we restricted the sample to women who had never previously undergone cervical cancer screening, findings may not transfer to WLHIV who have been screened before but decline repeat screening. The urban context, with many in self-employment (16/27, 59.3%), may underrepresent barriers in rural settings. Third, we did not collect data on ethnicity, religion, or clinical HIV characteristics, constraining exploration of cultural or clinical influences. Self-reported data may be subject to recall and social-desirability biases. Because barriers were elicited retrospectively in a single interview, participants’ accounts may reflect current reconstructions rather than the exact determinants operating at each prior declination episode.

Because one interviewer was clinic-affiliated and interviews occurred within the care setting, some participants may have moderated criticism or socially undesirable views despite assurances that participation was voluntary and independent of care. Because SEM levels were derived from WLHIV narratives only, findings reflect participant perceptions and may not capture provider or system viewpoints. We did not triangulate interview findings with provider interviews or additional clinical record review beyond screening-register verification. We also did not conduct participant checking, which may have limited opportunities for participants to refine or challenge our interpretations. Nearly half of participants could not estimate household income (13/27, 48.1%), which limited our ability to examine how financial barriers varied by income level, despite frequent cost narratives. We did not maintain field notes, which may have reduced contextual detail available during analysis. Because interviews sometimes prompted or coincided with emergent intention to screen, findings reflect a time-point snapshot and declining screening at the index visit should not be interpreted as durable refusal. Statements about earlier encounters are based on clinic screening registers and participant reports. Prior declinations were not observed prospectively. Register-documented prior declinations may not have fully distinguished active refusal from situational deferral or service disruption at every prior visit.

## CONCLUSIONS

Among WLHIV at an urban HIV clinic in Jos who declined cervical cancer screening at the index visit and at least one previous offer, perceived susceptibility to cervical cancer was often low or ambiguous despite high perceived severity. Although screening benefits were recognized, they were often outweighed by knowledge gaps and misinformation, emotional barriers including fear and anxiety about the procedure and possible results, and financial and logistical constraints, contributing to non-initiation of screening when it was offered. Guided by HBM and SEM, our findings highlight actionable levers within integrated HIV care, including fear reduction and stigma-sensitive counseling, predictable, streamlined clinic flow (shorter waits, reliable staffing), reduced indirect costs (transport support), and consistent provider invitations and reminders. Linking these institutional fixes with community-embedded messaging to address misinformation may improve uptake. Expressions of future willingness during interviews may indicate readiness to screen, although whether interview-expressed willingness translates into action remains uncertain. Future work should evaluate multilevel strategies tailored to this setting, including HPV self-sampling or screen-and-treat approaches where locally feasible, and should assess scalability beyond this single-site, urban context.

## AUTHOR CONTRIBUTIONS

**Conceptualization:** AA, PM, JM

**Methodology:** AA, SMI, PM, JM

**Investigation:** AA

**Formal analysis:** AA, SMI, PM, JM

**Project administration:** AA, MJA

**Supervision:** PM, JM

**Writing – original draft:** AA

**Writing – review & editing:** AA, SMI, MJA, AMK, YD, MGBN, PM, JM

## DATA AVAILABILITY STATEMENT

Because the data consist of in-depth qualitative interviews with women living with HIV and include sensitive, potentially identifying information, full transcripts cannot be publicly shared under the terms of ethics approval and participant consent. De-identified excerpts supporting the findings are included in the manuscript and tables. Requests for access to additional de-identified transcript excerpts may be directed to the Jos University Teaching Hospital Health Research Ethics Committee and will be subject to ethics approval and a data use agreement.

## ACKNOWLEDGMENTS

We are grateful to the women who participated in this study. We thank Grace C Job, Habila Augustina Daniel, Goodnews Oluwatosin Elijah, and Blessing Ishaya Ndacks at the AIDS Prevention Initiative in Nigeria (APIN) Clinic, Jos, for their assistance with participant recruitment and data collection.

## Notes

### Competing Interest Statement

The authors have declared no competing interest.

### Funding Statement

Research reported in this publication was supported by the Fogarty International Center of the National Institutes of Health under Award Number D43TW010540, through the Global Health Emerging Scholars program. The content is solely the responsibility of the authors and does not necessarily reflect the official views of the National Institutes of Health. The funder had no role in the design of the study, data collection, analysis, interpretation of data, or writing of the manuscript.

